# Genomic epidemiology of early SARS-CoV-2 transmission dynamics in Bangladesh

**DOI:** 10.1101/2024.05.07.24306987

**Authors:** L. Carnegie, J.T. McCrone, L. du Plessis, M. Hasan, M.Z. Ali, R. Begum, M.Z. Hassan, S. Islam, M.H. Rahman, A.S.M. Uddin, M.S. Sarker, T. Das, M. Hossain, M. Khan, M.H. Razu, A. Akram, S. Arina, E. Hoque, M.M.A. Molla, T. Nafisaa, P. Angra, A. Rambaut, S.T. Pullan, K.L Osman, M.A. Hoque, P. Biswas, M.S. Flora, J. Raghwani, G. Fournié, M.A. Samad, S.C. Hill

## Abstract

**Background:** Genomic epidemiology has helped reconstruct the global and regional movement of severe acute respiratory syndrome coronavirus 2 (SARS-CoV-2). However, there is still a lack of understanding of SARS-CoV-2 spread in some of the world’s least developed countries (LDCs).

**Methods:** To begin to address this disparity, we studied the transmission dynamics of the virus in Bangladesh during the country’s first COVID-19 wave by analysing case reports and whole-genome sequences from all eight divisions of the country.

**Results:** We detected >50 virus introductions to the country during the period, including during a period of national lockdown. Additionally, through discrete phylogeographic analyses, we identified that geographical distance and population -density and/or -size influenced virus spatial dispersal in Bangladesh.

**Conclusions:** Overall, this study expands our knowledge of SARS-CoV-2 genomic epidemiology in Bangladesh, shedding light on crucial transmission characteristics within the country, while also acknowledging resemblances and differences to patterns observed in other nations.

## 1. Background

Viral genomic analyses of severe acute respiratory syndrome coronavirus 2 (SARS-CoV-2) have provided insight into viral spread on both local and global scales and helped to inform infection control measures [1–6]. For instance, genomic analysis has been used to identify that Brazil shifted from being a net importer to an exporter of SARS-CoV-2 lineages over the course of 2020 [5], and to reconstruct patterns of within-country viral movements in France during the country’s first two Coronavirus disease 2019 (COVID-19) pandemic waves [7]. Using viral genomic data allows for epidemiological insight that cannot be captured from case data alone, including the inference of transmission dynamics prior to the first recorded sample or during periods with limited disease surveillance.

Despite often having high age-specific infection fatality rates (IFRs) [8], limited genomic studies of SARS-CoV-2 transmission dynamics have been conducted in the world’s least developed countries (LDCs) [8–10]. LDCs are low/middle-income countries (LMICs) that the United Nations (UN) classifies as possessing particularly high levels of poverty, human resource weakness, and economic vulnerability [10,11]. Instead, most studies focus on findings from high income countries (HICs), particularly in Western Europe and North America, likely reflecting a greater ability to invest in both sequence generation and research efforts [8,9,12]. This is problematic as findings from HICs may not be readily applicable to LDCs. LDCs often face unique challenges in controlling the virus, including limited public health resources, weak healthcare systems, and large sections of their populations highly susceptible to external economic pressures [8,9,13]. The indiscriminate application of control measures that are effective in high-income countries (HICs) may lead to unintended consequences in LDCs, particularly in settings where enforcement is challenging [13]. For example, stay-at-home orders have at times led to mass economic migration from cities, resulting in virus exports to larger geographic areas [14]. Identifying factors that influence transmission dynamics in LDCs allows for better tailoring of control measures to local epidemiological contexts, thereby increasing their chances of success [13,15].

The limited number of genomic epidemiological analyses in LDCs is exemplified in Bangladesh, where, despite some insights, many aspects of SARS-CoV-2 spread remain poorly understood. Cowley et al., (2021) compared observed mobility pattern data with time-scaled viral phylogenies to suggest that mass migration out of Dhaka, the capital of Bangladesh, may have driven early viral spread across the country [14]. However, in contrast to many HICs, the potential association between spatial and demographic factors and viral dispersal patterns has not been quantified [14]. Likewise, while studies reported that SARS-CoV-2 may have appeared in Bangladesh weeks before the first case report [14] and was likely introduced multiple times from several countries [16–18], the number of viral lineage imports to and exports from Bangladesh have not been estimated.

Bangladesh is the most densely populated country in the world that is not a city-state (165 million inhabitants living at over 1000 people/km^2^), and one of several LDCs in South Asia [14,19–23]. The first case of SARS-CoV-2 in Bangladesh was confirmed on March 8^th,^ 2020 [19,24]. In response to rising cases, a nationwide lockdown (known as a ‘National General Holiday’) was introduced on the 26^th^ of March and lasted until the 30^th^ of May. Restrictions and screening procedures were implemented at educational institutions, workplaces and international airports [23,25]. Infections nevertheless increased steeply in April, with nearly 313,000 confirmed cases reported by 31^st^ August 2020 (a total of 4,281 deaths) [18]. Following the end of the national lockdown, a subsequent zone-coded lockdown was used whereby movement within and between certain high-incidence areas was restricted [23,25–27].

This study aims to explore the spread of SARS-CoV-2 into and within Bangladesh during the first wave of the COVID-19 pandemic. We generated 175 SARS-CoV-2 whole-genome sequences from cases confirmed by molecular testing. These were combined with 140 additional sequences corresponding to confirmed cases occurring in Bangladesh during the study period to inform a phylodynamic analysis. We assessed the frequency and size of SARS-CoV-2 lineage imports to Bangladesh. We also identified drivers of viral dispersal among regions within the country, using a subset of 194 sequences from the countries’ two largest lineages.

## 2. Methods

### 2.1. Sample Collection

Samples were chosen for sequencing from residual anonymised diagnostic samples collected between 26^th^ April - 31^st^ August 2020 that were received by the National Reference Laboratory for Avian Influenza, Bangladesh Livestock Research Institute (BLRI) and that had tested positive by Real-time reverse transcription polymerase chain reaction (rRT-PCR). There was no patient or public involvement in the design of this study. Samples were nasopharyngeal or oropharyngeal swabs or both and were collected along the clinical criteria according to the National Guidelines on Clinical Management of Coronavirus Disease 2019 (COVID-19) published by the Directorate General of Health Services (DGHS), Bangladesh. A structured data collection questionnaire (Figure S1) was collected alongside all samples, detailing each patient’s geographic origin, clinical symptoms, travel history, and probable viral source. rRT-PCR for SARS-CoV-2 detection was performed using fluorescent probes and the result was considered positive when the cycle threshold (Ct) values of both the ORF1ab and N genes were <32 and <35, as measured by ROX [28] and FAM [29] dyes [30–32]. We attempted to choose samples proportionally to confirmed cases per district (see Table S1 for a comparison of sequences to confirmed cases per district), whilst also still including a high percentage of samples from likely under-sampled regions. We preferentially selected samples for which information on travel history (including confirmation of absence of travel) was present.

### 2.2 Sequencing

We sequenced 175 whole SARS-CoV-2 genomes from samples using a Nanopore GridION and the nCoV-2019 sequencing protocol v3 (LoCost) V.3 incorporating the ARTIC multiplex-PCR sequencing pipeline (as described in [33]). We combined newly generated sequences with 140 Bangladeshi sequences accessed (July 2022) via the GISAID [34] EpiCOV database (www.gisaid.org) collected from patients during the same sampling period. These included: (i) sequences already used in published papers (n = 139) and (ii) unpublished sequences where submitting laboratories granted us permission for their inclusion (n = 1). All SARS-CoV-2 sequences were aligned using Llama (Local Lineage and Monophyly Assessment) [35] v0.1 with default parameters. Using AliView [36] v1.28 we confirmed that there were no sequences from the alignment that either were duplicated, short (<70 % of the total sequence length), or indicative of containing sequencing or assembly errors.

### 2.3. Epidemiological Data

We obtained COVID-19 confirmed case data between 26^th^ April - 31^st^ August 2020 from the Institute of Epidemiology Disease Control and Research (IEDCR) “COVID-19 Dynamic Dashboard for Bangladesh” [37]. Aggregated daily case counts were available for each of Bangladesh’s eight divisions (Chattogram, Dhaka, Sylhet, Khulna, Barishal, Rajshahi, Rangpur, Mymensingh). For the national-level data, days in which data was missing from one or more divisions were excluded (n = 10) from analyses.

### 2.4. Epidemiological Analyses

We applied methods implemented in EpiFilter [38] to estimate the median and 95% equal-tailed Bayesian credible intervals of the effective reproduction number at time *t*, (R(t)), in each division between 17^th^ April 2020 and 31^st^ August 2020. As detailed in [38], this method reduces the mean squared error in R(t) inference by applying optimal recursive smoothing. We used the SARS-CoV-2 serial interval distribution detailed in Ferguson et al. (2020) as the generation time distribution [39]. We also use a weekly averaging filter to minimise the impacts of inconsistent reporting on R(t) inference.

### 2.5. Transmission Lineage Analyses

We first assembled a global alignment by sampling SARS-CoV-2 genome sequences evenly for each week and country (oldest sequence: 2019-12-24, newest sequence: 2020-08-31). Temporal outliers, defined as samples that fell beyond 4 interquartile ranges of the expected divergence given were identified and removed using TreeTime [40] and a fixed clock rate of 0.00075 substitutions/site/year. This resulted in a dataset of 5804 sequences. We then used ThorneyBEAST (https://beast.community/thorney_beast) to estimate a posterior molecular clock tree using a pipeline similar to that of du Plessis et al., (2021) [41], Raghwani et al., (2022) [4] and Gutierrez et al., (2021) [42]. This employed a strict molecular clock model [43] (0.00075 substitutions/site/year), and a Skygrid coalescent tree prior [44]. For the starting tree, we used a maximum-likelihood tree produced with IQ-TREE [45] under a Jukes-Cantor substitution model, using Wuhan/WH04/2020 as an outgroup. During the Markov chain Monte Carlo (MCMC), we constrained our tree search to sample only node height and resolutions of polytomies present in the starting tree. We completed 5 chains of 200 million steps with sampling every 1.8 million steps, removing the first 10% of steps as burn-in. The resulting sample of 500 trees from the posterior were used to generate a maximum clade credibility (MCC) tree. We confirmed that the Effective Sample Size (ESS) of all parameters was >200 using Tracer [46] v1.7.1.

We then conducted a discrete trait analysis (DTA) with these 500 empirical trees in BEAST [47] to estimate ancestral node locations (Bangladesh or non-Bangladesh). We used a robust counting approach [48] to evaluate the expected number of location state transitions into and out of Bangladesh across a posterior sample of 2000 trees.

Finally, we used the location-annotated MCC tree to identify transmission lineages in Bangladesh, following previously described methods [41]. Briefly, nodes with a posterior probability > 0.5 inferred to be located in Bangladesh are considered “Bangladeshi nodes”. A depth-first search initiated at a random Bangladeshi node is run until complete or until a non-Bangladeshi node is encountered, with encountered nodes labelled as belonging to the same transmission lineage. A new starting node is chosen until every Bangladeshi node is visited, and hence all Bangladeshi tips have been assigned a lineage or classified as a singleton. For each Bangladeshi lineage, we estimated time to the most recent common ancestor (TMRCA).

### 2.6. Discrete Trait Analysis with Generalized Linear Model

To investigate how viral dispersal was influenced by spatial and demographic factors in Bangladesh we focused on the two largest transmission lineages identified in the global dataset (Material and Methods Section 2.5.) Only those whole-genome sequences with known sampling districts were used (n = 142/307 for lineage 2 and n = 52/97 for lineage 8, remaining total with sampling districts = 194) (Tables S2-3). To facilitate effective discrete phylogeography analyses, we reduced the original 28 districts (second administrative level), covering six out of the eight Bangladeshi divisions (first administrative level) into a smaller set of discrete locations. In brief, we employed k-means clustering based on Euclidean distance, as implemented in the ’*stats*’ package in R [49] v4.1.2, to group these sequences into eight geographic regions, each representing a grouping of several districts. All sequences were assigned to their respective district centroid coordinates for this purpose. We assembled data on chosen predictors for each of these eight geographical groups of districts: population size and density, the ratio between the number of cases and population size, and geographic distance (ellipsoid distance between grouping centroids) (see Tables S4-5 for details). Whilst we did not expect a strong temporal signal given the low mutation rate of SARS-CoV-2 and the short timescale of our study (e.g., [14,50]), we used TempEst [51] v1.5.3 to check that the temporal signal in each lineage alignment was not incompatible with using fixed substitution rates based on the literature and to check for outlier sequences (Figure S2). We used a DTA model implemented in BEAST [47] v1.10.4 to reconstruct virus lineage movements between the eight pre-defined geographic groupings. A strict molecular clock model with a fixed rate (0.001 substitutions/site/year) and an SRD06 substitutional model [52] was shared between both transmission lineages to improve convergence, with a separate exponential growth coalescent tree prior that allows for different timings of lineage growth. We used a faster clock rate (0.001 substitutions/site/year) in these shorter timescale phylogeographic analyses, compared to the previous global transmission lineages analyses (0.00075 substitutions/site/year), to account for the existence of slightly deleterious mutations circulating in the Bangladesh transmission lineages that had not been removed by purifying selection [53]. We used separate generalised-linear models (GLMs) to determine which covariate(s) (Table S4-5) best predicted the frequency of viral lineage movements between locations for each lineage. The migration rates in the DTA model are parameterised as a GLM using covariates as coefficients of the GLM. DTA-GLM analyses that included either population density or population size predictors were run separately because of the multi-collinearity between these predictors. For each analysis, we executed two chains of 150 million steps logged every 15,000 steps, removing the first 10% of steps as burn-in. We used Tracer [46] v1.7.1 to confirm that the ESS was >200 for all parameters. The support of the Bayes Factor (BF) for transmission between discrete traits was interpreted as described previously in [54], with BF > 100 ‘very strong support’, and BF > 10 indicating ‘strong support’.

## 3. Results

### 3.1. Epidemiological Findings

COVID-19 confirmed cases increased towards the end of the national lockdown and continued rising during the subsequent zone-coded lockdown, peaking at the start of July 2020 (Figure 1). Reported case numbers dropped sharply during Eid al-Adha, which is one of the most important Muslim festivals and marked by a period of public holiday in Bangladesh [55] (31^st^ July 2020 – 1^st^ August 2020) (Figure 1). This is consistent with reduced reporting and testing in other countries in periods surrounding important religious events [56].

**Figure 1.**
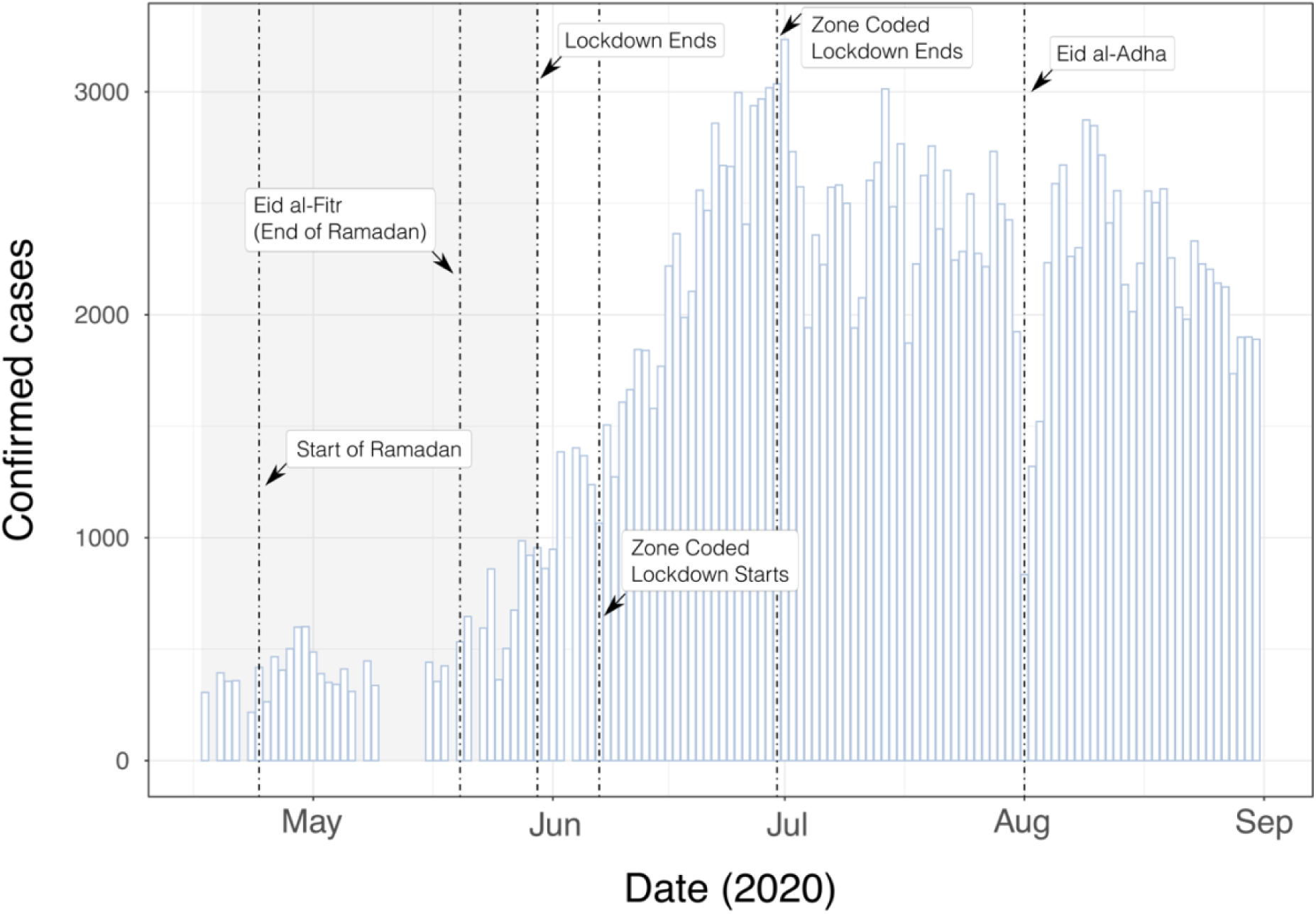
Daily COVID-19 confirmed case counts in Bangladesh, 2020. Grey highlighted region indicates the timing of a national lockdown. The x-axis ticks correspond to the start of the named month.

We plotted daily COVID-19 confirmed case counts in Bangladesh’s eight divisions between 17^th^ April 2020 and 31^st^ August 2020. Case counts in each division were similar to the pattern observed at the country level (Figures 1 & 2A). It is possible that rising cases could be somewhat driven by increased testing capacity over time [14]. In particular, during the first months of the pandemic, residents in Dhaka, the country’s largest and most populated city, had easier access to SARS-CoV-2 testing compared to other regions in Bangladesh [14,57]. Consequently, the Dhaka division reported the majority (50.5%) of confirmed SARS-CoV-2 cases in the country (Figure 2A) [14,57]. Spikes in reported cases during the first wave of the pandemic were observed in some other divisions, e.g., in Rangpur in the start of July 2020 (Figure 2A).

**Figure 2.**
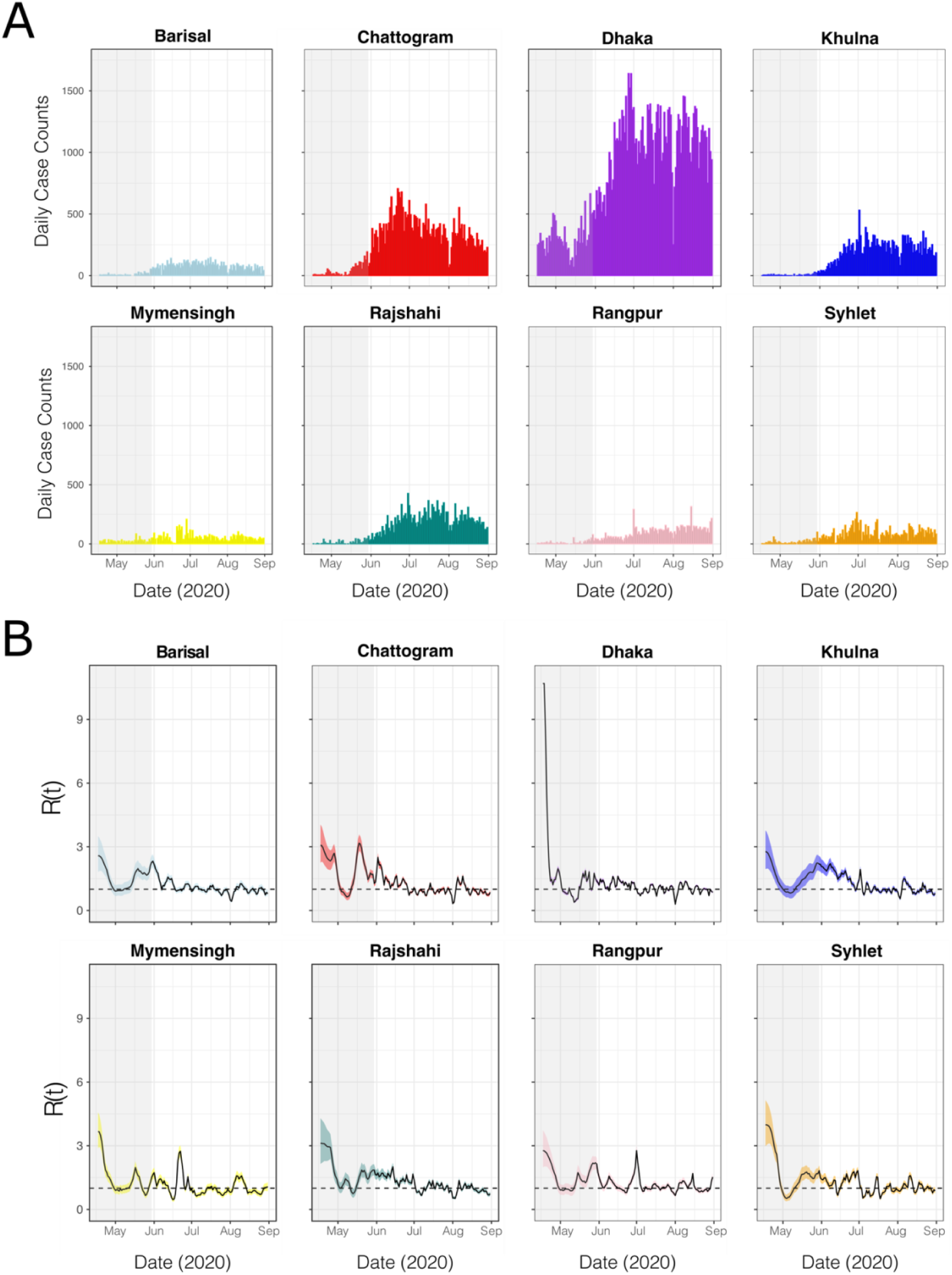
Grey-highlighted regions indicate a national lockdown. A) Daily COVID-19 confirmed cases in each division. B) Effective reproduction number (R(t)) by division. Median Rt estimates are shown by black lines, and 95% Bayesian credible intervals by coloured ribbons.

Division-specific effective reproduction numbers R(t) showed similar temporal trends in most Bangladeshi divisions (Figure 2B). R(t) typically fell in the first half of the national lockdown, but then peaked above 1 near the middle or end of May, concurrent with both the religious festival of Eid al-Fitr and the end of the national lockdown. R(t) then slowly decreased and fluctuated around 1 by the start of July (Figure 2B). Division-specific spikes and troughs in R(t) were also observed, such as a reduction of R(t) below 1 in mid-June in Mymensingh (Figure 2B).

### 3.2. Transmission Lineage Findings

We estimated the number of SARS-CoV-2 lineage importations over time to Bangladesh. The estimated number of viral lineage movements into the country (n = 111, 95% HPD: 99 – 121) was approximately 4 times higher than the number of viral lineage movements from Bangladesh to other countries globally (n = 26, 95% HPD: 16 – 38) (Figure 3A), indicating that Bangladesh was mainly an importer of virus lineages during the first wave.

**Figure 3.**
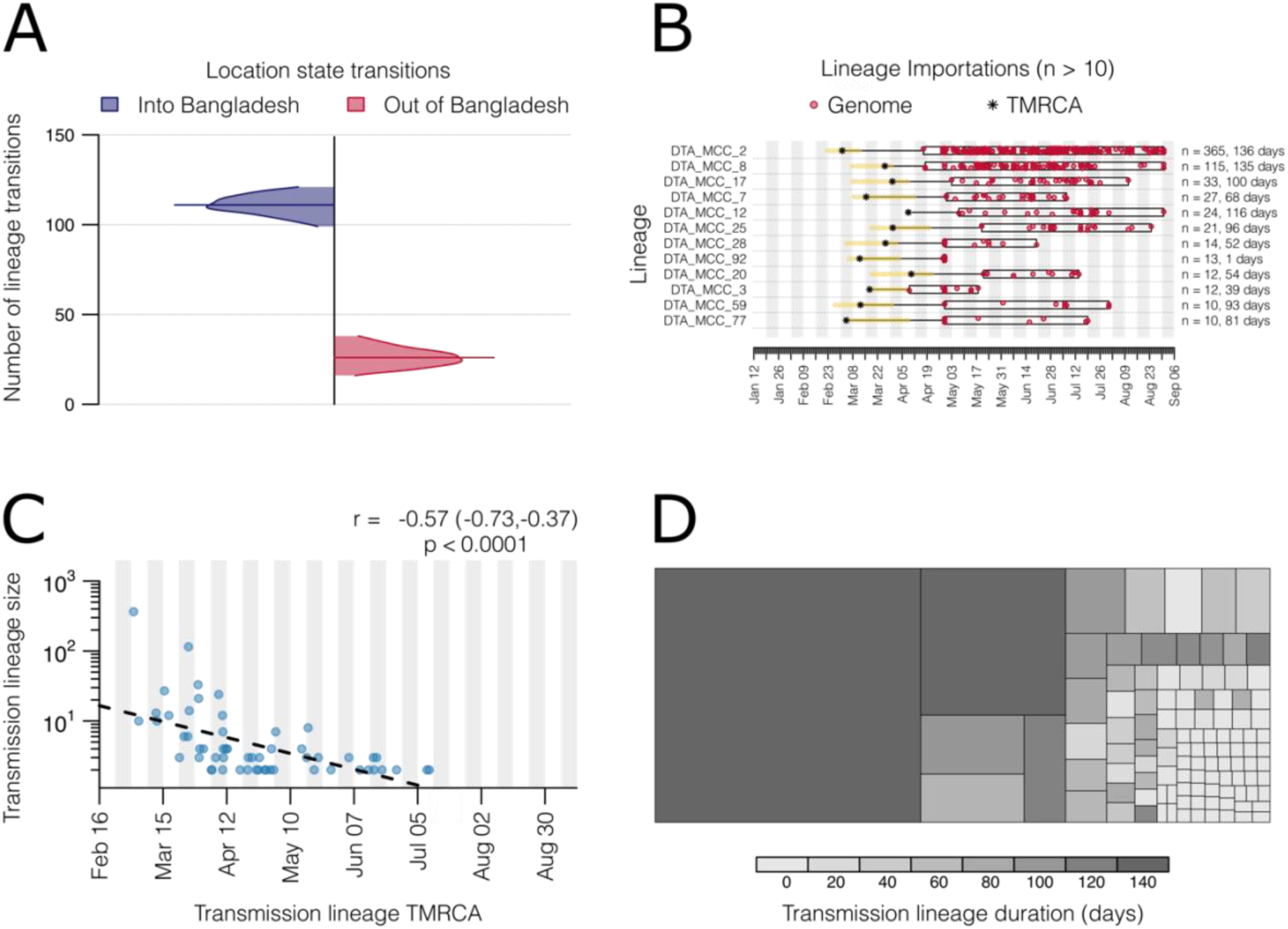
SARS-CoV-2 transmission lineage characteristics in Bangladesh. A) Number of location state transitions between the phylogenetic traits Bangladesh/Global (imports into Bangladesh = blue, exports from Bangladesh = red), as detected via the robust counting approach. Posterior distributions are truncated at their 95% highest posterior distribution (HPD) interval limits and median estimates are shown using horizontal lines. B) Duration and timing of the largest Bangladesh transmission lineages (> 10 genomes). Each row represents a transmission lineage, and red dots indicate genome sampling times. Boxes and labels on the right axis show the sampling duration (see Figure S3 for more details on sampling duration per lineage), and number of sampled genomes (n). Asterisks show the median estimated time to most recent common ancestor (TMRCA) for each lineage, with the 95% HPD as a yellow bar C) Relationship between transmission lineage size and TMRCA, with a dashed line indicating the slope of a linear regression. The Pearson correlation coefficient, 95% confidence interval, and p-value are shown. D) Cells represent transmission lineages and singletons, each coloured by estimated duration and scaled by size.

We detected 56 distinct Bangladesh transmission lineages (i.e., lineages spreading within Bangladesh, represented by >1 genome), and 51 singletons (singular Bangladesh genomes representing lineage imports originating from non-Bangladesh regions, yet without evidence of autochthonous transmission) (Figure S3). Twelve lineages contained >10 sequences, and two lineages contained >100 genomes. Several lineages (n= 16, 28.6%), including the two largest lineages, had an estimated TMRCA that fell either prior to or within the following week of the start of the national lockdown on the 26^th^ March (Figures 3B & S3) [25]. Multiple lineages, however, had TMRCAs overlapping with the period of nationwide lockdown (Figures 3C & S3). Finally, there was a strong negative exponential relationship between the estimated TMRCA and transmission lineage size (Figure 3C). Larger transmission lineages in Bangladesh were generally associated with longer sampling durations (Figure 3D).

### 3.3. Drivers of virus spread within Bangladesh

We used DTA models coupled with GLMs to identify significant predictors of virus lineage dissemination among eight geographical groups each containing districts from multiple Bangladeshi divisions (Figure 4A), using only those sequences from the two largest Bangladesh transmission lineages (Figure 3). We fitted two models with either population size or population density as the demographic predictor (covariate values shown in Table S5 & Figure S4). We observed consistent directionality and effect sizes for two population metrics in each of their respective models, but notable differences were found in the BF support and inclusion probabilities. In the model including population density as a predictor (Figure 4B), there was strong evidence (BF > 10, following: [54]) that virus lineages were more likely to move from areas with high population density. We also found very strong support (BF > 100) for virus movement between geographically close locations and very strong support (BF > 100) towards areas with a high number of confirmed cases per capita. In the model including population size as predictor (Figure 4C), there was strong support for virus spread occurring more commonly to and from areas with high population sizes, and for virus movement towards locations with a high number of confirmed cases per capita. However, distance was not identified as a significant predictor (BF < 1).

**Figure 4.**
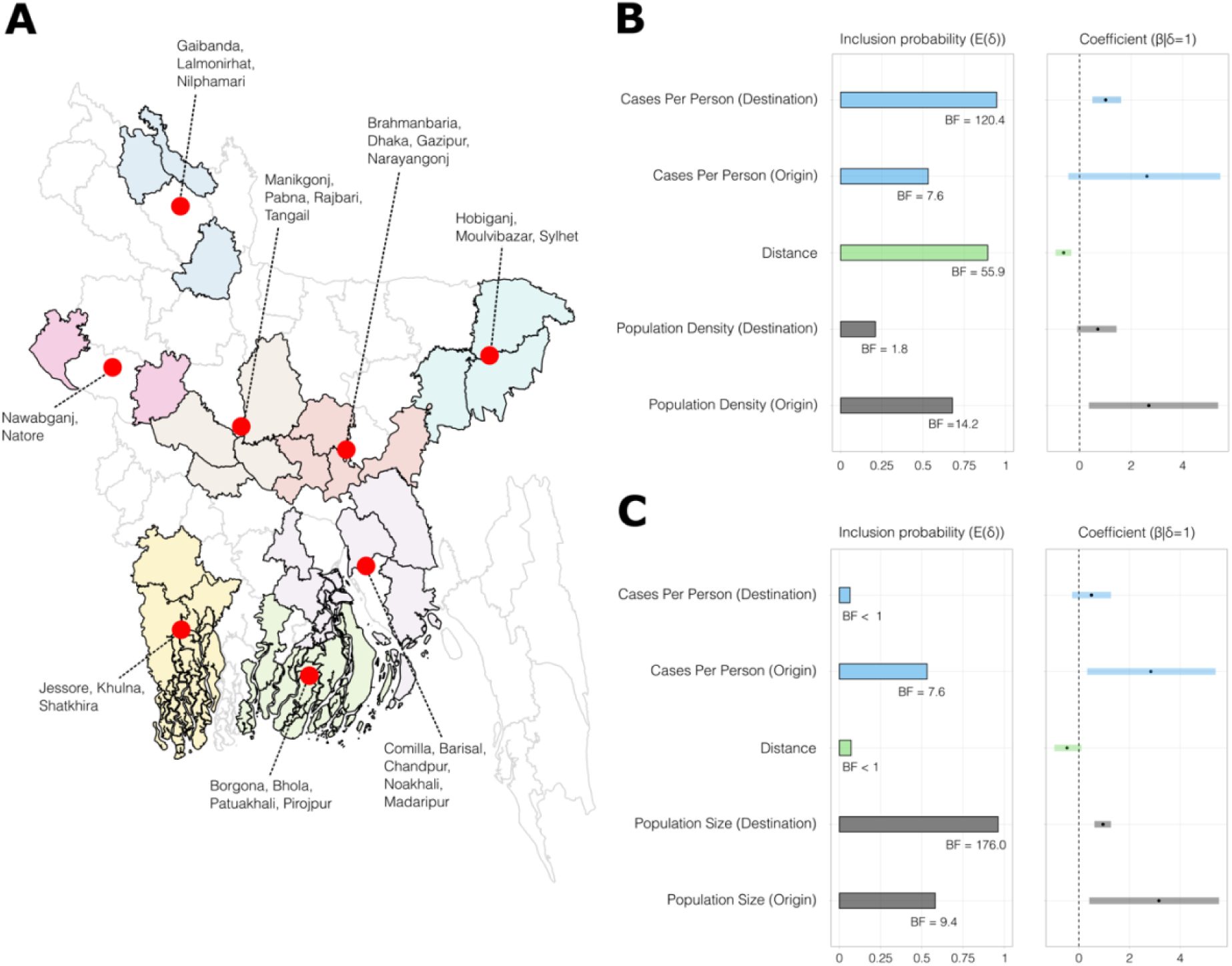
Factors associated with SARS-CoV-2 spread. A) Map of Bangladesh showing the eight geographical groups of districts used as discrete traits in the DTA-GLM. Region centroids are marked by red dots. B and C) Predictors of SARS-CoV-2 spread based on models with either population density (B) or population size (C) included as predictors. Bar and line colours indicate different covariates, with origin and destination predictor of a covariate given the same colour within each plot. Inclusion probability is the posterior expectation that the indicator variable is associated with each predictor E(δ) and suggests that the predictor is associated with different rates of viral diffusion. Bayes Factor (BF) support values for each covariate are indicated by black text annotations. The coefficient (β|δ=1) represents the contribution of each predictor on a log scale when the predictor is included in the model, with the 95% credible interval of the GLM coefficients (β) represented by horizontal lines.

**Figure 5.**
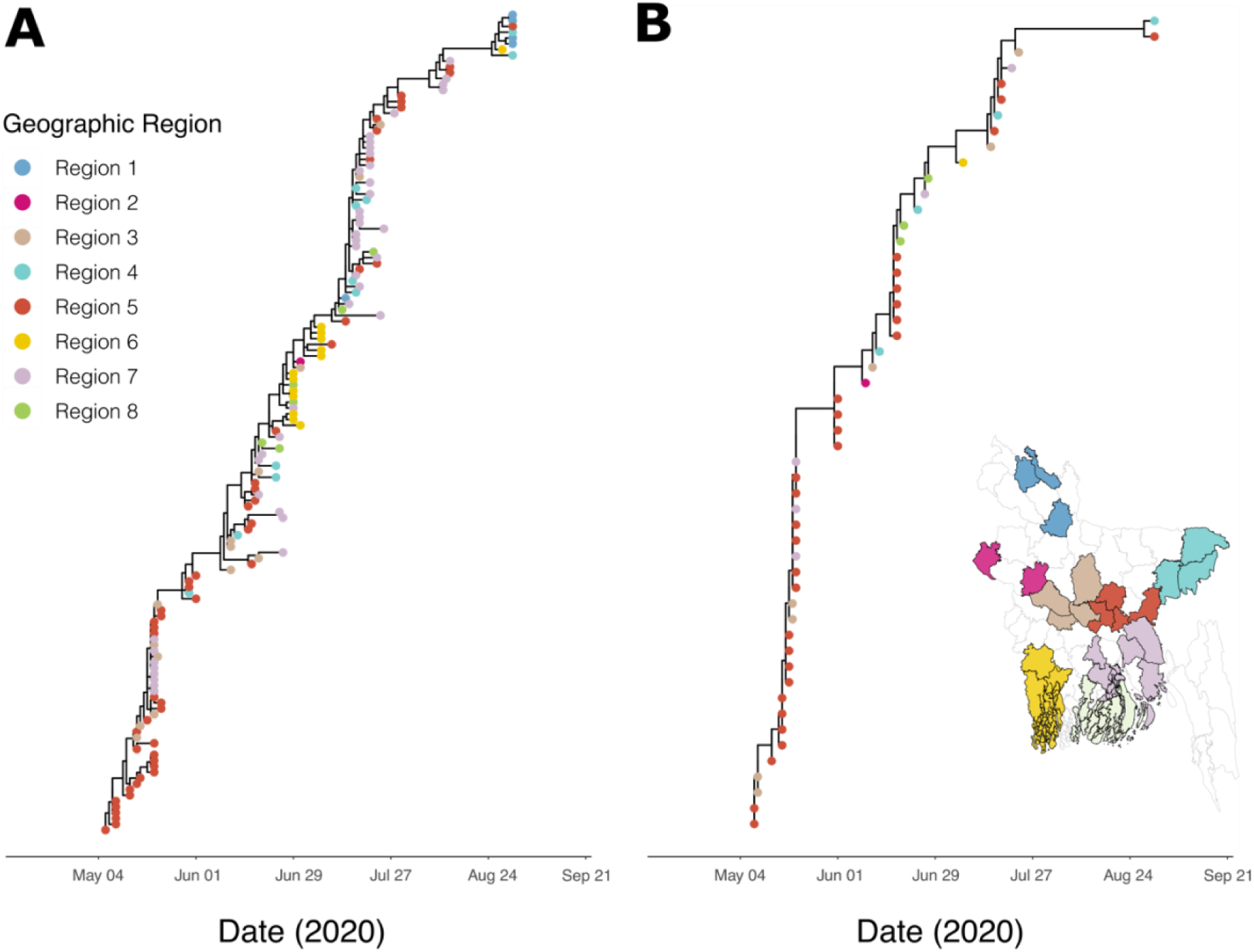
MCC phylogenies of the two largest lineages detected. A) Lineage 2, and B) Lineage 8. Branch lengths represent time, as shown on the axis. Tips are coloured by the sampling geographic region used in the DTA-GLM analyses, as shown in the inset map.

## 4. Discussion

Genomic epidemiology has played a crucial role in understanding the global and regional movement of SARS-CoV-2. However, research efforts have disproportionately neglected studying virus spread within the world’s least developed countries. To contribute to addressing this disparity, we use a combination of epidemiological and phylodynamic analyses to investigate the transmission dynamics of SARS-CoV-2 in Bangladesh during the country’s initial wave of COVID-19. We determined that R(t) showed a declining trend during the start of the national lockdown but rose slightly towards the end of the national lockdown in late May 2020, which was perhaps driven by activity surrounding Eid al-Fitr. Subsequently, R(t) slowly stabilised and fluctuated around ∼1 in all divisions. Our study revealed that cases in Bangladesh were initiated by more than 50 instances of virus introduction into the country, with multiple lineages maintained even during the period of national lockdown. Finally, we identified using discrete phylogeographic analyses that patterns of virus dispersal may have been shaped by population density and/or size, as well as geographical proximity, although the latter was not consistently important across all analyses.

The detection of over 50 virus introductions during Bangladesh’s initial COVID-19 wave, and of multiple persistent transmission lineages during the lockdown, contrasts with studies from Hong Kong [58] and New Zealand [59], where virus cases were linked to repeated individual introductions rather than persistently present lineages. Both our study and the Hong Kong study were based on similar pipelines adapted from du Plessis et al. (2021) [41], and therefore the observed difference is likely not caused by different study designs. Instead, the distinction likely reflects a greater inability to adequately maintain social distancing in Bangladesh’s high-density urban areas and the relatively greater capacity of these other countries to control local transmission through measures such as community surveillance and contact tracing [12,44]. Additionally, New Zealand and Hong Kong adopted stringent inbound travel regulations and testing measures [58,59]. In contrast, Bangladesh had limited thermal scanners at Dhaka international airports, and reported low quarantine compliance [19,60]. These differences may have played a further role in the observed variations in SARS-CoV-2 lineage dynamics between locations.

In congruence with previous studies in South Africa [6] and Italy [61], we found that the estimated number of virus importations into the country considerably outweighed exports. The return of a large number of Bangladeshi nationals to the country during the early stages of the pandemic may have allowed for extensive lineage importation to the country from a wide range of geographical locations [14,16–18,62]. Notably, over 10 million Bangladeshi citizens live abroad, and the number of migrant labourers returning to the country was 8 times higher in 2020 than in previous years [18,63,64]. Introductions may have been seeded via international airports [14,16–18,60], as well as through cross-border human mobility with neighbouring countries (e.g., India [65–67]). The latter has been an important pathway for the introduction of SARS-CoV-2 lineages to other LDCs (e.g., Burkina Faso [68] and Rwanda [69]). However, as the number and origin of introductions and exports estimated from genomic data are dependent on sample size [41], our analyses may be biased towards capturing imports rather than exports, given relatively less intense sampling in Bangladesh compared to many other regions globally.

Viral lineages moved more frequently from locations with higher to locations with lower population density or total population size. This finding may reflect the spread of viral lineages across the country during the early stages of the pandemic being driven by mass migration out of major cities/urban hotspots with relatively higher case rates to rural areas, following the announcement or extension of the national lockdown, as previously suggested by several publications that reference news reports (e.g., [19,70]) and a comparative analyses of mobility patterns with genomic data [14]. The considerable number of commuters from peripheral rural locations into urban areas may also have contributed to virus spread following the ease of the national lockdown [14,71–73]. This pattern of wide-scale movement from urban areas driving early spread of the virus has previously been identified using genomic methods in both HICs (e.g., France [7]) and regions of lower-middle-income countries (LMICs) (e.g., Gujarat, India [4]). As such, in conjunction with this previous research, our study’s findings may further support the notion that population size/density and mass human movements were generally important drivers of within-country spread during this stage of the pandemic, regardless of country wealth [74].

In one of our discrete phylogeographic analyses, we identified that viral movement occurred preferentially between more proximate geographic groupings of districts in Bangladesh. This suggests that certain lockdown measures (e.g., travel advisories, railway closures, domestic flight suspensions [23,25–27]), likely had a more substantial impact on restricting long-distance human mobility and virus spread, compared to movement and viral dispersal between closer or neighbouring divisions. Distance has also been identified as a predictor of SARS-CoV-2 spread in several other countries, including Israel and Brazil [75,76]. That said, the presence of strong evidence (using BF support) for distance depended here on whether human population density or size was considered, with only strong evidence detected when the population density predictor was also included in the model. This could indicate that distance had a relatively weak effect on viral transmission dispersal between regions in the country, perhaps because Bangladesh is relatively small and its population is highly mobile [72].

Our study has several limitations. Firstly, only a very small fraction of the total confirmed SARS-CoV-2 cases could be sequenced due to resource constraints. Greater sampling and sequencing of cases would have allowed us to analyse the virus dispersal patterns in Bangladesh at higher resolution. Second, the limited genomic surveillance in certain locations outside of Dhaka where many cases were confirmed, particularly the absence of sequences from Chattogram, the country’s second-largest city, indicates that our analyses may have underestimated the extent and frequency of virus movements within Bangladesh. As such, the outputs of our DTA-GLM models may have not fully captured the strength or impact that certain demographic predictors had on within-country viral dispersal [14]. We attempted to mitigate this limitation by choosing samples for sequencing roughly proportionally to the confirmed cases per Bangladeshi district while retaining most samples from any particularly under sampled locations. However, for several regions, very few samples were available and case reporting is likely to be inherently spatially biased. Third, due to the short time-period of our study, we were unable to investigate changes in the drivers of viral spread during the study period. The dynamics of transmission and the sources of infection may have altered over time, especially in response to mobility restrictions and interventions such as border closures. Furthermore, the geographic origin of each sequence was determined based on individuals voluntarily disclosing their present place of residence, which can be prone to inaccuracies. Likewise, missing location metadata meant we had to reduce the number of sequences from each lineage to only sequences with known division when performing the DTA-GLM phylogeographic analyses, which may have reduced the possible statistical power of the analyses.

## 5. Conclusions

In conclusion, this study on SARS-CoV-2 genomic epidemiology during the first wave in Bangladesh uncovers distinct and common virus transmission patterns in comparison to other countries of various income levels. These findings highlight the significance of genomic epidemiological analyses in resource-constrained regions like Bangladesh in potentially helping to inform the design of more specific and accurate evidence-based interventions aimed at reducing the import and within-country spread of viral outbreaks. Addressing the limitations in genomic surveillance in Bangladesh is crucial for improving the capacity to respond to future viral outbreaks.

## Data Availability

The datasets and BEAST XML files supporting the conclusions of this article are available in the Github repository. The dataset for the newly reported genetic sequence data supporting the conclusions of this article are available in GISAID EpiFlu database (https://gisaid.org) under the accession numbers included within the additional files of this article [Table S6].

https://github.com/lorcancarnegie/SARS_CoV_2_Bangladesh

## List of abbreviations

BF: Bayes Factor
COVID-19: Coronavirus disease 2019
Ct: cycle threshold
DTA: discrete trait analysis
ESS: Effective Sample Size
GLMs: generalised-linear models
HICs: high income countries
LDCs: least developed countries
LMICs: low/middle-income countries
MCC tree: maximum clade credibility tree.
MCMC: Markov chain Monte Carlo
rRT-PCR: real-time reverse transcription polymerase chain reaction
SARS-CoV-2: severe acute respiratory syndrome coronavirus 2
TMRCA: time to the most recent common ancestor
UN: United Nations

## Declarations

### Ethics approval and consent to participate

Patient consent for publication: Not applicable.

### Ethics approval

The study was carried out under the ethical approval from the ethical approval committee of Bangladesh Livestock Research Institute, Bangladesh with the reference number BLRI/EA/2020102/2022, and with informed consent from tested individuals.

### Consent for publication

Not applicable.

### Availability of data and materials

The datasets and BEAST XML files supporting the conclusions of this article are available in the ‘SARS_CoV_2_Bangladesh”repository, [https://github.com/lorcancarnegie/SARS_CoV_2_Bangladesh]. The dataset for the newly reported genetic sequence data supporting the conclusions of this article are available in GISAID EpiFlu database (https://gisaid.org) under the accession numbers included within the additional files of this article [Table S6].

### Authors’ contributions

L.C., S.C.H, G.F. and J.R. conceived the phylodynamic and epidemiological analysis. L.C. wrote the manuscript and performed the phylodynamic and epidemiological analysis. S.C.H, J.R. and G.F. assisted with phylodynamic and epidemiological analysis. JT.M. and L.D.P. performed the viral lineage analyses. S.C.H., G.F. and J.R. edited the manuscript and supervised the work. M.A.S, M.H., M.S.S., and S.I. contributed to the collection of epidemiological data, laboratory testing, and generation of sequences and helped with interpreting the findings. All other authors contributed to fieldwork or laboratory work design or implementation. All authors gave final approval for publication.

## Acknowledgements

We thank the Child Health Research Foundation (CHRF), Bangladesh, for allowing us to use their data on the GISAID EpiCoV database (accession numbers in Table S7-8). We also acknowledge the use of additional sequences from the GISAID EpiCoV database for the global dataset used in our lineage analyses and provide accession numbers and acknowledgements in the supplementary electronic material (Table S7-8). We thank the Directorate General of Health Services (DGHS) and Institute of Epidemiology Disease Control and Research (IEDCR) Bangladesh, DGHS, Ministry of Health and Family Welfare for sharing COVID-19 samples and data.

## Footnotes

S.C.H and M.S are joint last authors.

The findings and conclusions in this report are those of the author(s) and do not necessarily represent the official position of the U.S. Centers for Disease Control and Prevention.

