## Supplementary Appendix for "Genomic epidemiology of early SARS-CoV-2 transmission dynamics in Bangladesh"

**Institute of Epidemiology, Disease Control and Research (IEDCR)**

**COVID-19Suspected Case RecordForm**

IEDCR

IDNo: Date:……./………../2020

| | | | |
|---|---|---|---|
| **Institute/Hospital:** | | | |
| Department: | | | |
| Unit: | Unit Head: | | |
| Ward: | Ward/Cabin: | | |
| **Name of Patient:** | | | |
| Age: | …….Years or if <5 years ……….. Month | Sex: | □Male □Female |
| **Occupation:** | | Phone/Cell No: | |
| **Address:** | | Emergency Contact Number | |
| **Email address:** | | | |
| **Referred by:** | | | |

**COVID-19 Suspect Criteria: Give (√) to select**

| Symptom | Yes/No | |
|---|---|---|
| 1. Fever (≥38°C or100.4°F) | □**Yes** | □**No** |
| 2. Headache | □**Yes** | □**No** |
| 3. Cough | □**Yes** | □**No** |
| 4. Breathlessness | □**Yes** | □**No** |
| **5.** Others (Specify) | | |

6. Clinical or radiological evidence of Pneumonia or severe Acute Respiratory Distress Syndrome □**Yes** □**No** □**Unknown**
7. **Travel history** in 14 days before illness onset. □**Yes** □**No** □**Unknown**
   If yes, **Country**……………………..
   **Date of departure** from the place……………………………………….
8. Has the person **had contact with a confirmed case** in the 14 days prior to symptom onset?
   □**Yes** □**No** □**Unknown**
9. Has the person **visited any health care facility** in the 14 days prior to symptom onset?
   □**Yes** □**No** □**Unknown**

**Concurrent risk factors (Check all that apply):** □COPD □Asthma □ Interstitial lung disease □DM □IHD □HTN □CKD □CLD □Malignant disease □On steroid therapy □Pregnancy □Others ………………………………………………

**Specimen:** □Collected □ Not collected, if collected mention type:
□Nasal swab □ throat swab □Sputum □Tracheal aspirate □Serum
□Other:……………………………………………………………………

**If any remarks:**

Interview Conducted by

**Figure S1.** Questionnaire used to obtain epidemiological information from patients alongside swab samples.

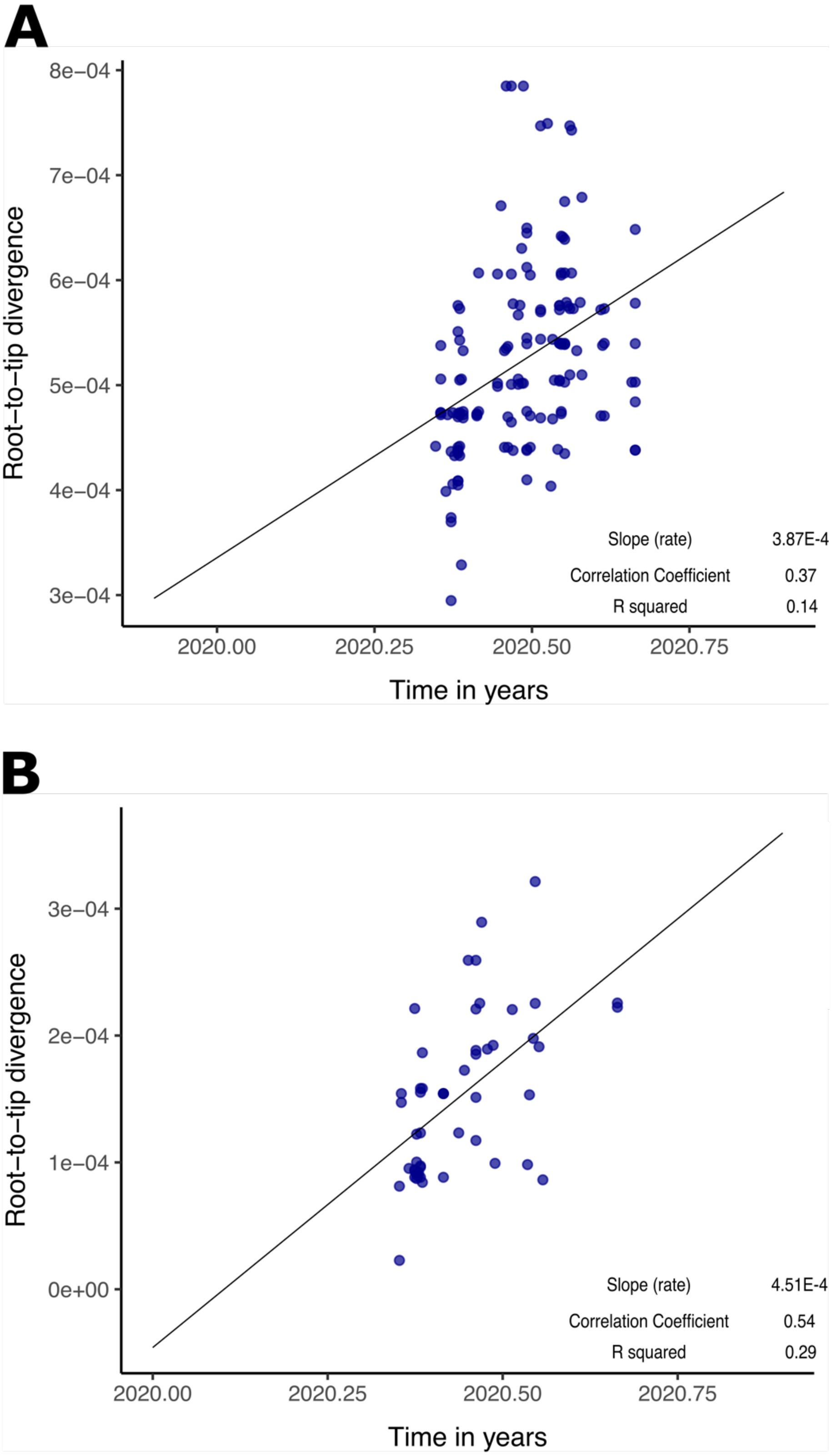


***Figure S2.*** *Root-to-tip divergence plots and the associated statistics for the **A**) lineage 2 dataset, and the **B**) lineage 8 dataset.*

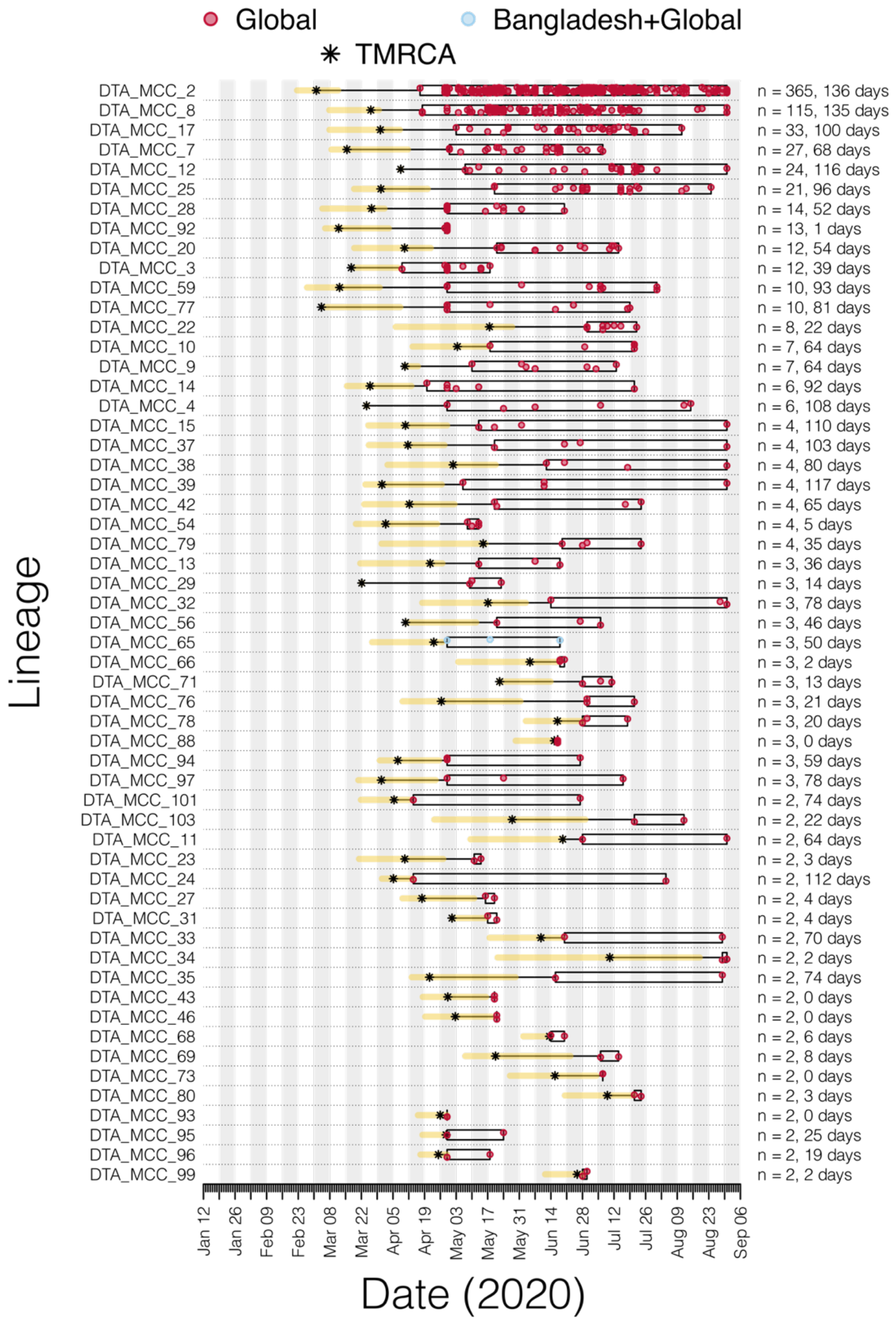


***Figure S3.*** *Duration and timing of Bangladesh transmission lineages. Each row represents a transmission lineage, and red dots indicate genome sampling times. Boxes and labels on the right axis show the sampling duration, and number of sampled genomes (n). Asterisks show the median estimated TMRCA of each lineage, with the 95% HPD as a yellow bar.*

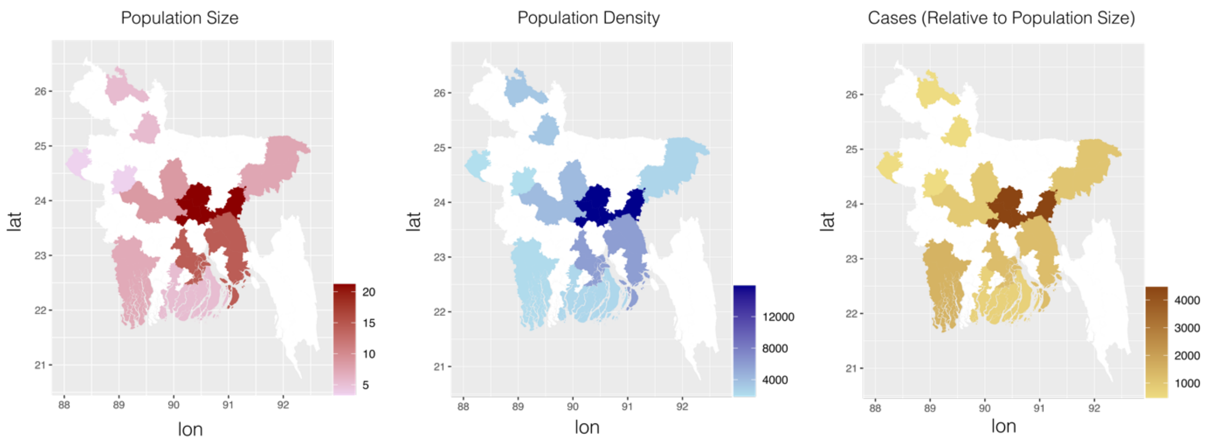


***Figure S4.*** *Choropleth maps for each predictor in the DTA-GLM. Values for each district on the map represent the mean value for the geographic region it belongs to. Regions not represented in this study are in white.*

***Table S1.*** *Total number of sequences per district compared to number of confirmed cases during the sampling timeframe.*

| District | Number of Sequences | Number of Confirmed Cases in Study Period |
|---|---|---|
| Gaibanda | 3 | 1057 |
| Lalmonirhat | 1 | 734 |
| Nilphamari | 1 | 826 |
| Nawabganj | 1 | 677 |
| Natore | 1 | 893 |
| Manikgonj | 14 | 1354 |
| Pabna | 5 | 1424 |
| Rajbari | 1 | 2791 |
| Tangail | 1 | 2422 |
| Hobiganj | 4 | 1224 |
| Moulvibazar | 9 | 1524 |
| Sylhet | 2 | 5130 |
| Brahmanbaria | 12 | 2315 |

| | | |
|---|---|---|
| Dhaka | 48 | 84048 |
| Gazipur | 9 | 3308 |
| Narayangonj | 13 | 5260 |
| Jessore | 6 | 3600 |
| Khulna | 5 | 6147 |
| Shatkhira | 3 | 1024 |
| Comilla | 8 | 6148 |
| Barisal | 3 | 3548 |
| Chandpur | 15 | 2426 |
| Noakhali | 17 | 4406 |
| Madaripur | 3 | 1467 |
| Borgona | 1 | 954 |
| Bhola | 3 | 752 |
| Patuakhali | 3 | 1156 |
| Pirojpur | 2 | 1090 |

***Table S2.*** *List of districts in each geographical grouping.*

| **Geographical Region** | **District** |
|---|---|
| Region 1 | Gaibanda |
| Region 1 | Lalmonirhat |
| Region 1 | Nilphamari |
| Region 2 | Nawabganj |
| Region 2 | Natore |
| Region 3 | Manikgonj |
| Region 3 | Pabna |
| Region 3 | Rajbari |
| Region 3 | Tangail |
| Region 4 | Hobiganj |
| Region 4 | Moulvibazar |

| | |
|---|---|
| Region 4 | Sylhet |
| Region 5 | Brahmanbaria |
| Region 5 | Dhaka |
| Region 5 | Gazipur |
| Region 5 | Narayangonj |
| Region 6 | Jessore |
| Region 6 | Khulna |
| Region 6 | Shatkhira |
| Region 7 | Comilla |
| Region 7 | Barisal |
| Region 7 | Chandpur |
| Region 7 | Noakhali |
| Region 7 | Madaripur |
| Region 8 | Borgona |
| Region 8 | Bhola |
| Region 8 | Patuakhali |
| Region 8 | Pirojpur |

***Table S3.*** *Number of sequences per geographic grouping (total, and per lineage).*

| Geographical Region | No. Seq. | No. Lineage 2 Seq. | No. Lineage 8 Seq. |
|---|---|---|---|
| Region 1 | 5 | 5 | 0 |
| Region 2 | 2 | 1 | 1 |
| Region 3 | 21 | 14 | 7 |
| Region 4 | 15 | 11 | 4 |
| Region 5 | 82 | 51 | 31 |
| Region 6 | 14 | 13 | 1 |
| Region 7 | 46 | 41 | 5 |
| Region 8 | 9 | 6 | 3 |

***Table S4.*** *GLM Covariate details and sources*

| Covariate | Data Source |
|---|---|
| Population Size (Millions) | Bangladesh Population and Housing Census 2011: http://www.bbs.gov.bd/site/page/47856ad0-7e1c-4aab-bd78-892733bc06eb/Population-and-Housing-Census |
| Population Density (100m$^2$) | We determined the total area of each region in km$^2$ using the area function of raster package. We then calculated the population density of each region by dividing the total area by the total population size (Bangladesh Population and Housing Census 2011: http://www.bbs.gov.bd/site/page/47856ad0-7e1c-4aab-bd78-892733bc06eb/Population-and-Housing-Census /). We finally multiplied by 10000 to give values greater than 1, giving a value for population density on a 100m$^2$ scale. |
| Mean Daily Confirmed Cases (per Million People) | Institute of Epidemiology Disease Control and Research (IEDCR) "COVID-19 Dynamic Dashboard for Bangladesh, hosted at http://103.247.238.92/webportal/pages/covid19.php# <br><br> We eliminated days where data was missing from any region, and then calculated the mean daily case count for whole study period. We then divided this mean value by the total population size of the region (Bangladesh Population and Housing Census 2011: http://www.bbs.gov.bd/site/page/47856ad0-7e1c-4aab-bd78-892733bc06eb/Population-and-Housing-Census /). |

***Table S5.*** *GLM predictor values for each geographical region (group of divisions).*

| Geographical Region | Population Size (Millions) | Population Density (100m$^2$) | Mean Daily Confirmed Cases (Per Million People) |
|---|---|---|---|
| Region 1 | 5.47 | 10.80 | 3.77 |
| Region 2 | 3.36 | 9.22 | 4.77 |
| Region 3 | 8.57 | 10.25 | 7.34 |
| Region 4 | 7.44 | 8.53 | 7.79 |
| Region 5 | 21.23 | 36.01 | 32.64 |
| Region 6 | 7.07 | 7.51 | 12.09 |
| Region 7 | 14.41 | 13.40 | 9.12 |
| Region 8 | 5.32 | 7.75 | 5.71 |

***Table S6***. *GISAID EPI SET acknowledgements table for 175 newly published sequences* **(see separate file).**

***Table S7***. *GISAID EPI SET acknowledgements table for 103 sequences from lineage 2 accessed from* GISAID [34] epiCOV database (www.gisaid.org) **(see separate file).**

***Table S8***. *GISAID EPI SET acknowledgements table for 37 sequences from lineage 8 accessed from* GISAID [34] epiCOV database (www.gisaid.org) **(see separate file).**
